# TrajectoryViz: Interactive visualization of treatment trajectories

**DOI:** 10.1101/2024.04.01.24305168

**Authors:** Maarja Pajusalu, Kerli Mooses, Marek Oja, Sirli Tamm, Markus Haug, Raivo Kolde

**Author notes:** Corresponding author: Maarja Pajusalu Postal address: Arvutiteaduse instituut, Tartu Ülikool Narva mnt 18 51009 Tartu Estonia.

## Abstract

**Background and objectives:** With the proliferation of real-world or observational health data, there is increasing interest in studying treatment trajectories. The real-life treatment trajectories can be complex, and one has to simplify the patterns to draw any conclusions; however, oversimplification will cause the loss of essential details. Thus, the visualization challenge is to strike a balance between the two extremes.

**Methods:** We have implemented the observation of treatment trajectories starting from cohort definitions in cooperation with medical specialists, data processing, and then generating the interactive visualizations and detailed data tables within an open-source R package as a Shiny dashboard. The created R package called TrajectoryViz (https://github.com/HealthInformaticsUT/TrajectoryViz) enables reproducible visual analysis and visual content generation for various data investigations.

**Results:** We illustrate the use of the tool by assessing the sequence of events present within the data of cervical cancer prevention pathways, as well as the proportions of timely follow-up procedure events.

**Conclusion:** Building a toolset to access, manage, and analyze observational health data enables more accessible visual analysis of complicated data, adding time dimension to otherwise simplified event sequences that make up trajectories.

## 1 Introduction

The amount of routinely collected real-world health data (RWD) is rapidly increasing. The data stored in healthcare claims and electronic health records (EHR) is an invaluable source of information that can be used for a broad spectrum of research [1]. The analysis and visualization of such data have a great potential to discover new associations between the disease and previous or following health conditions and treatment patterns. Moreover, it can be used to describe how patients are treated in real life and assess their adherence to clinical guidelines. Clinical guidelines are systematically developed evidence-based recommendations trying to balance the benefits and harms of the treatment for specific medical conditions. As both under- and overtreatment can pose serious health risks to the patient and also a burden to the healthcare system, these guidelines must be followed. We aim to visualize treatment trajectories to identify what clinical events occur before and after certain events.

Visualizing treatment trajectories with RWD comprises several challenges as the data is complex in velocity, variety, and volume [2]. This means that traditional two-dimensional static charts cannot convey all the information stored in RWD. Currently, the advanced visualization techniques for large and complex EHR datasets are limited [3], and treatment trajectories are often visualized through the sunburst diagram, which resembles a multi-layer pie chart depicting the proportions of varying event sequences [4]. In the case of medical data, this enables to see the proportions of the cohorts receiving specific treatments or procedures at consecutive levels or layers. While giving an excellent overview of cohort division between different treatment patterns, the sunburst diagram lacks details like lengths of treatment periods, gaps, lengths of gaps between consecutive treatments, etc. Therefore, there is a need for improved, dynamic, and interactive visualization tools that allow the exploration of various data elements and facets simultaneously and manipulate visual elements [2, 3, 5].

To address this shortcoming, we have developed an R package called TrajectoryViz (https://github.com/HealthInformaticsUT/TrajectoryViz), which gives a detailed overview of the patient-level paths and different sequences. Our goal was to create a tool with an easy-to-use interactive interface that supports visual explorational analysis of sequence-based data as patients’ treatment trajectories. The main objective was to enable zooming into different subsections of the data under observation and visualizing the timespan between events on the selected trajectories. The package provides the researchers with graphical outputs from their analysis and tables with input data processed into various details, i.e., collecting and counting the frequencies of all unique trajectories or all individual trajectories with each event labeled with its sequence number on that path.

The paper aims to give an overview of the TrajectoryViz package and to illustrate its use through a use case. In the Methods section, we provide an overview of the package and the data pre-processing for executing the TrajectoryViz package. In addition, we describe a case study. In the Results section, we will explain the package’s functionalities using the case study as an example.

## 2 Methods

### 2.1 The Overview of the Package

TrajectoryViz is an R package that can visualize trajectories based on discrete or continuous treatment periods with the following data structure: SUBJECT_ID, STATE_LABEL (or STATE, both are accepted), STATE_START_DATE and STATE_END_DATE. The state label represents the values of any trajectory - these may be events, diagnoses, procedures, or any other value used to generate color coding for the trajectory visualization. The execution function (“trajectoryViz()”) creates an interactive R Shiny dashboard to read the input data, shows a data overview - number of subjects and unique paths - and displays an interactive sunburst plot of the proportions of unique sequences. Selecting a specific segment from the sunburst diagram (i.e., all sequences where event B follows event A) will plot out patient-level visualizations of all the patients with the treatment pathway starting with these events. The patient-level sequences are plotted as one color-coded line per patient with gray gaps between events, then sorted and aligned by selected events on the third plot on the dashboard. Finally, the package creates a funnel visualization to give a simple drill-down of subject counts on every filtering layer, starting from the number of subjects in the original data set to the number of patients, with the detailed conditions defined by selections made from the plots. All these visualizations are interactive, allowing both quantifying the interesting aspects or zooming into patterns when selecting more detailed sequences.

The analysis is not constrained to any particular data type, like treatment or diagnosis. Any kind of sequential data (column STATE_LABEL or STATE both are accepted) can be visualized. The user can observe, filter out, and download the detailed data tables generated on the second tab.

TrajectoryViz is designed to work with health data transferred to the Observational Medical Outcome Partnership common data model (OMOP CDM) [6], which means that the user could benefit from using an online tool ATLAS [7] for defining the cohorts. To make the visualization compatible with any OMOP formatted database, TrajectoryViz relies on the Cohort2Trajectory package in R [8, 17]. The cohort2Trajectory package summarizes multiple cohorts defined in ATLAS into linear simplified event sequences, which TrajectoryViz requires as input. Figure 1 illustrates the process from the database to the dashboard. At the same time, the TrajectoryViz package can be still used if the data is not in OMOP CDM format as long as the input CSV file has the minimal structure needed: SUBJECT_ID, STATE_LABEL (or STATE), STATE_START_DATE and STATE_END_DATE.

**Figure 1.**
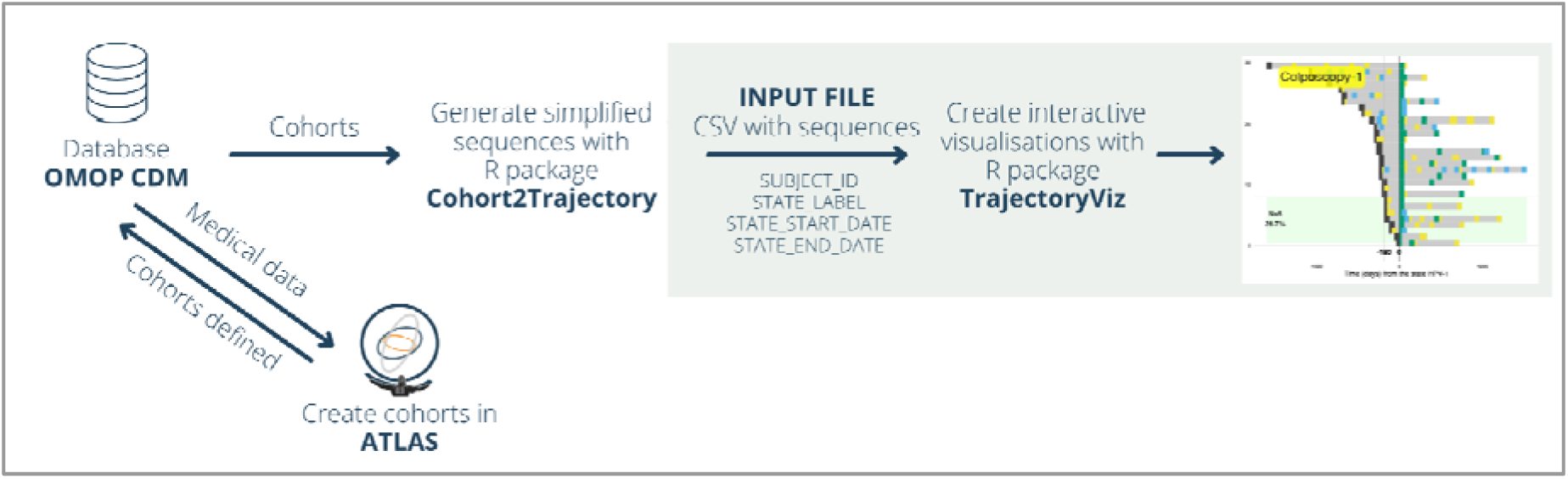
The pre-process for TrajectoryViz.

### 2.2 Case Study

To illustrate the capabilities of the TrajectoryViz package, we focus on cervical cancer screening with a PAP test. Cervical cancer is a common malignant neoplasm among women, which is preventable with timely discovery [9, 10]. Moreover, 12% of invasive cervical cancer cases are attributable to insufficient follow-up after receiving abnormal PAP tests [11]. Therefore, it is of utmost importance to provide effective and on-time treatment. In the case of slightly abnormal cytologic findings, the follow-up activities are less invasive and often need a repetition of a PAP test after a specific period. As for more severe high-grade cytologic findings, the need for additional tests is more urgent. The current case study focuses on cases where the PAP test resulted in high-grade squamous intraepithelial lesion (HSIL). According to the guidelines, this pathology requires an additional diagnostic procedure, colposcopy, within three to six months. We visualize the follow-up procedures of the first HSIL diagnosis to evaluate the adherence to national guidelines for diagnosing, monitoring, and treatment of precancerous cervical, vaginal, and vulvar conditions.

For this purpose, we used the data from Estonian EHR and healthcare service provision claims from 01.01.2015 to 31.12.2019. These two databases have been previously linked and transformed to OMOP CDM [12], allowing the ATLAS online tool to generate cohorts. The main cohort consisted of women with HSIL (N = 176) with the inclusion criteria: 1) female aged 30-60, 2) the first occurrence of HSIL after 01.01.2015 (index date), 3) no occurrence of more serious cervical pathology within 30 days before and after the index date, 4) no previous diagnosis of the corresponding pathology within three years before the index date, 5) no history of cervical cancer, hysterectomy, HIV, 6) no current pregnancy 7) a follow-up period at least 12 months.

To assess the occurrence and sequence of PAP tests, HPV tests and colposcopy following and preceding the cervical pathologies, we created additional cohorts for 1) PAP test, 2) HPV test, and 3) colposcopy, including all corresponding events from 01.01.2015 to 31.12.2019.

We summarized the data from all cohorts into simplified event sequences using the Cohort2Trajectory package [8]. In the case of timely overlap, we present events with higher priority. We prioritized the cohorts, starting from the highest priority: Colposcopy, HSIL, PAP, and HPV.

Through the visualization of this data, we aim to find answers to the following questions:

1. What is the proportion of women who receive colposcopy within six months after the HSIL and, thus, comply with the guidelines?
2. Are there additional PAP and HPV tests after the HSIL and before the colposcopy?
3. What are the following events after the first colposcopy?

The questions are answered within the results section and illustrated in Figures 3 and 4.

## 3 Results

### 3.1 The data

We initiate the package from RStudio with the function “trajectoryViz()” and upload the prepared CSV file. Uploading the data will generate the first visualizations and tabulated information on the sequences imported. An overview of uploaded data will be displayed, showing the number of patients and unique sequences in the data set (Figure 2, left side). The user can cut the data to start all sequences from a specific state. The sequences with a selected state not on the 1st level (first in sequence) will be cut from the start until the state chosen becomes the first. Cutting can be beneficial to simplify the visualizations. On the other hand, this means that the cut trajectories will lose some states before the selected event. The case visualized on the lower right sunburst in Figure 2 shows precisely that - now the trajectories do not show the event of “HSIL” result while all these events happened before the 1st Colposcopy.

**Figure 2.**
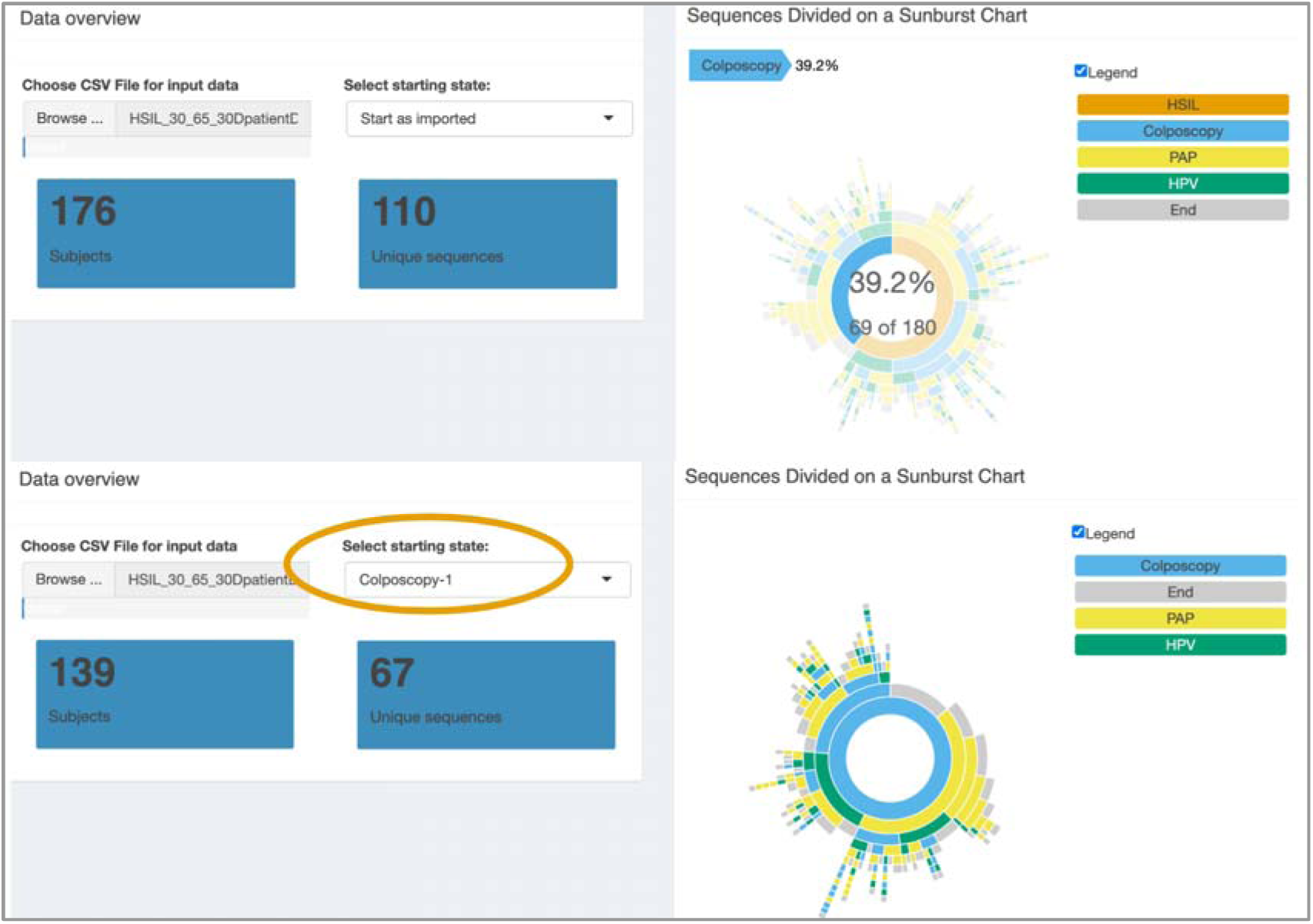
The uploaded data set (176 subjects and 110 unique sequences) was visualized on a sunburst diagram - upper part. A total number of sequences (176) is rounded up on the sunburst visualization to 180, while the function used there has native rounding. Lower part - the uploaded data set cut from 1st occurrence of Colposcopy (139 subjects and 67 unique sequences left).

### 3.2 The sunburst plot – the proportional division between sequences

The first visualization generated after data upload is a sunburst plot (Figure 2, right side) of treatment pathways showing the first-level state (a treatment, diagnosis, test, or any other event) in the center and subsequent events on the outer layers. Hovering any segment will indicate the corresponding number of individual trajectories following that sequence and their percentage of all trajectories imported. It will also highlight the state labels on that sequence. This functionality is depicted in Figure 2, top right side, showing 69 sequences and 39,2% of all starting with colposcopy. Each color represents a state (event, procedure, test, etc.). The sunburst is clickable to visualize the treatment pathways of the segment clicked on. If the sunburst has a full circle on the inner-most layer, all sequences start with the same event, and the proceeding plot will show all trajectories of the data set.

### 3.3 Temporal visualization of individual treatment pathways

To concentrate on the patients with HSIL measurement on their pathway, we can click on the inner- most HSIL segment on the sunburst diagram (Figure 2, top). The package generates a plot that visualizes each treatment pathway on a separate row of color-coded sequences (Figure 3, left side). Each color represents a state (event, procedure, test, etc.), and the length of the color indicates the consecutive period in days the patient had this state on their pathway. In the case of a procedure, it would most often be just a short bar on the plot (1 day), whereas, in the case of prescriptions covering a more extended period, the colored bar might be longer. The states have a sequence count added to the state label - first, “HSIL” is now labeled “HSIL-1” while there are also repetitive events; as an example, the fourth colposcopy procedure for the same subject is labeled “Colposcopy-4”.

**Figure 3.**
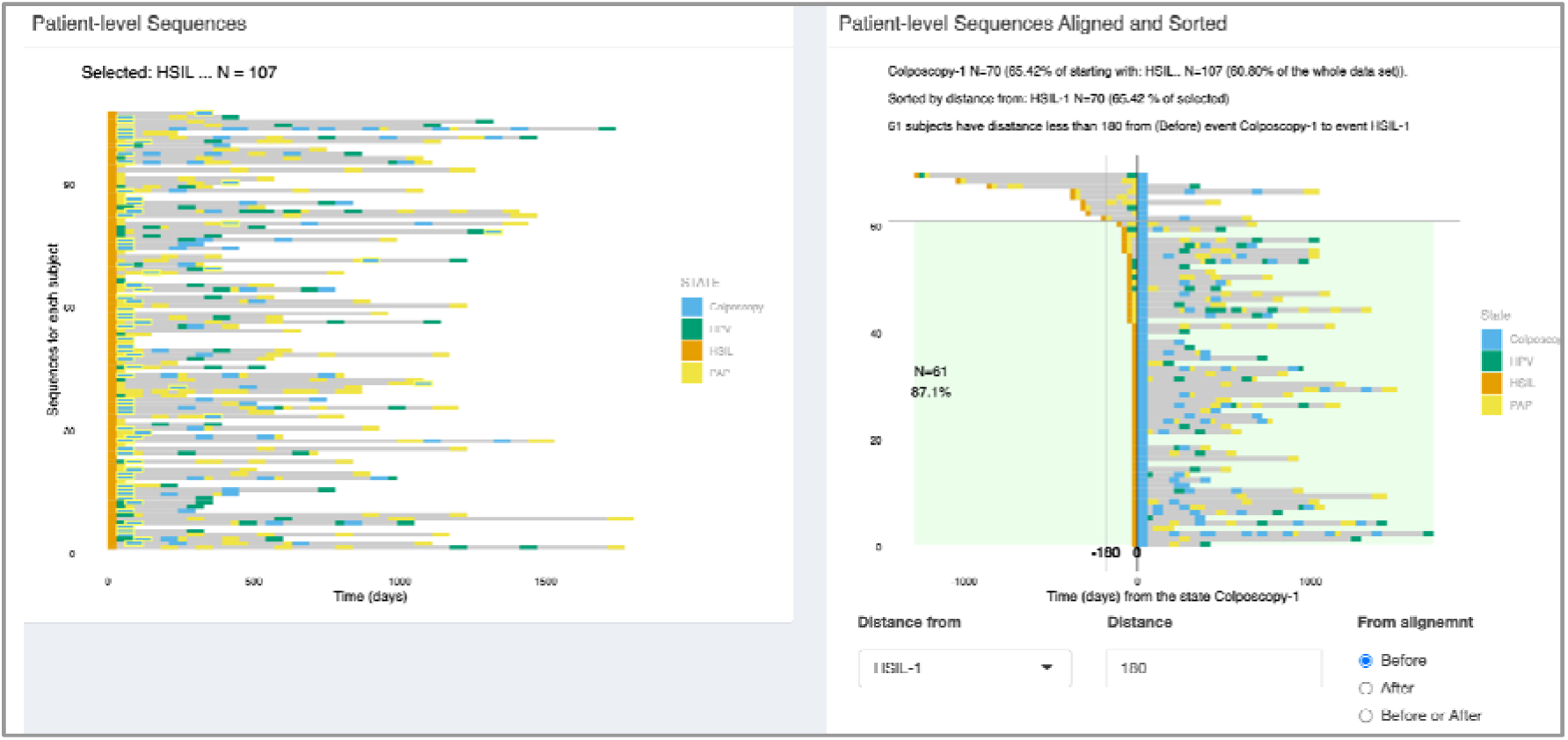
The selected subsample of 107 patients and their treatment events are sequenced out (left side). We are highlighting the “Colposcopy-1” event to be aligned. The plot on the right aligns the trajectories by the distance between the alignment event (Colposcopy-1”) and sorts by the time difference from the selected event (“HSIL-1”). The green area indicates the cohort with a time difference of fewer than 180 days in our case study.

The plot is clickable to align and sort the sequences on the following plot. Clicking on a state on any line generates a time-aligned and sorted visualization of selected pathways. In our case study, we already selected patients with HSIL and wanted to study the timeline of the following colposcopy procedure. We see (Figure 3) that 107 patients in the selected subsample had an event of “HSIL” on their treatment trajectory. It must be declared, though, that having an event of “Colposcopy” already states that the subjects in our cohort must have had a corresponding test result. Still, it could have also been either before entering the timeframe of our dataset, or it was overridden by the more prominent (selected by ourselves in data preparation) state, in our case, the “Colposcopy.” In this case, the patients do not have “HSIL” events in our data and are not visualized within that subsample.

### 3.4 Aligned and sorted temporal visualization of selected pathways

According to the treatment guidelines, colposcopy should be performed within three to six months after the “HSIL” diagnosis. We can adjust the plot by:

1. Selecting the state from which the distance will be calculated for sorting.
2. Adding the timeframe indicator in time units (days).
3. Selecting either before or after the alignment event.

The resulting plot will visualize the sorted pathways with the number and percentage of subjects falling into the selected timeframe.

On the right side of Figure 3, we can see the answer to our case study question 1 - a visualization after aligning the sequences by the procedure “Colposcopy-1”, sorting by the distance from “HSIL-1” and indicating that we are interested in the sequences with a time difference of fewer than 180 days (6 months) before Colposcopy-1. The light green area shows the valid trajectories - 61 patients (87% of current selection) who met the treatment guidelines and did experience the procedure within a suitable time frame.

We answer our case study question 2 on the same plot (Figure 3, right side). According to the treatment guidelines, there should not have been new events such as “PAP” or “HPV” between “HSIL” and “Colposcopy,” but we can see all the yellow and green events that appear in the top area of the plot.

As we were also trying to find answers to our questions about the timing of events following the colposcopy (case study question 3), we cut the original data set from “Colposcopy-1”, resulting in 139 subjects (Figure 2, below). We generate the plot by clicking on the first state on any of the lines to see a plot where the pathways are aligned by colposcopy and reordered by the time distance from another event. As an example, we first reorder the plot by the time distance from the state “PAP-1” (Figure 4, left side) and then from “Colposcopy-2” (Figure 4, right side) to observe the events and time gaps after “Colposcopy-1”.

**Figure 4.**
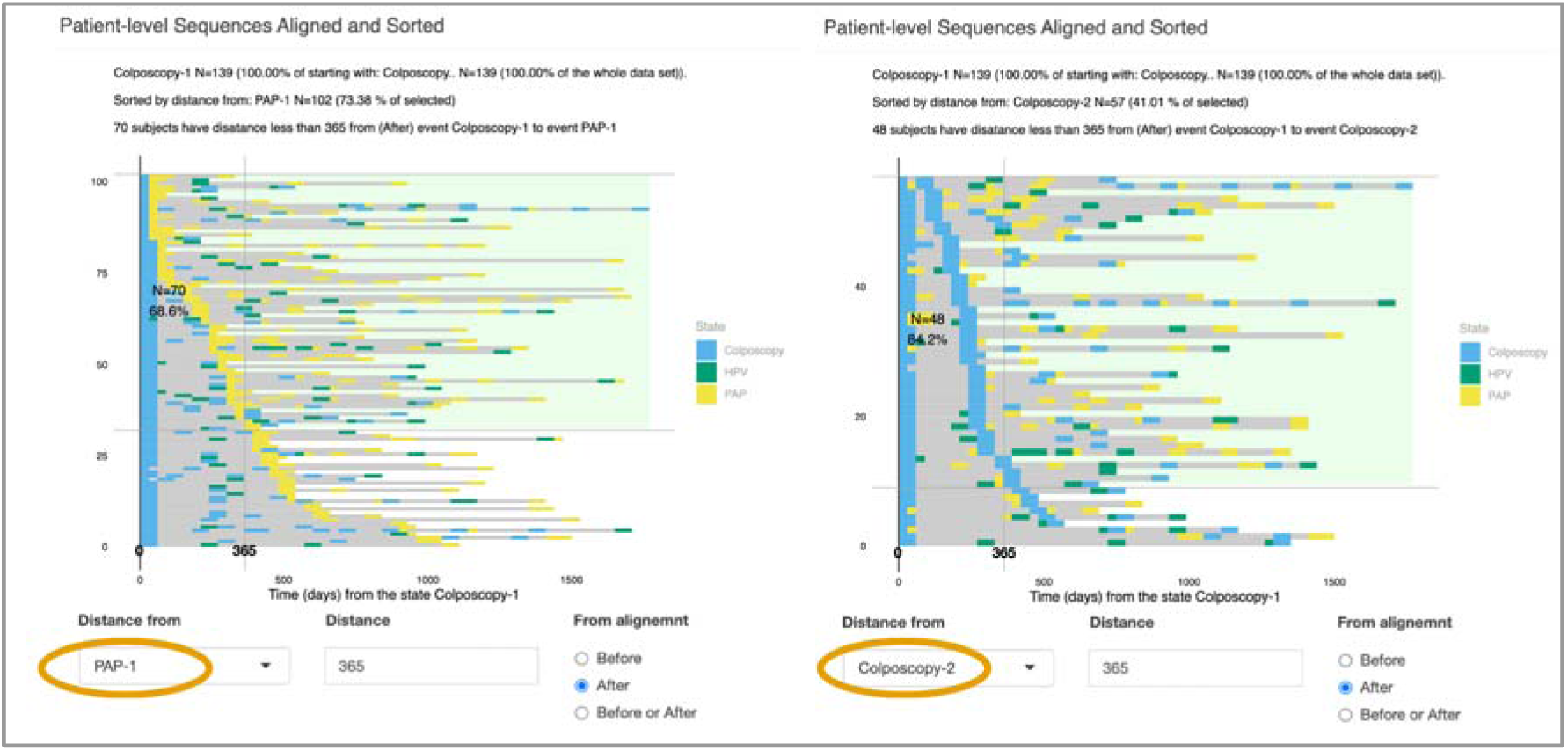
Observing the events after the first Colposcopy. Example of treatment trajectories starting with and aligned by Colposcopy, sorted by distance from events PAP (in yellow, on the left) and Colposcopy-2 (in blue, on the right).

The temporally aligned and sorted view also informs the percentage of filtered sequences that did meet the criteria for the time gap between selected events. We chose a time frame of 365 days after the first event to see how many patients had the follow-up PAP test or recurring colposcopy procedure within that time. To see these numbers more visually and be able to use the visual for studies and articles, we have added a funnel visualization (Figure 5) that shows a drill-down of the number of subjects on each filtered level. In our example, we started with 176 subjects, from which 107 were filtered out as starting with “HSIL”; from those, 70 subjects did have the procedure “Colposcopy” present. As we searched for the distance from “HSIL”, no one was filtered out during the sorting process, and finally, 61 subjects of the subgroup were compliant with the time measurement of 180 days between the events. Figure 5 shows the funnel, and on the right side, the interactivity of the visualization is also shown. By hovering over the mouse, we can see percentages of each level compared to all preceding levels.

**Figure 5.**
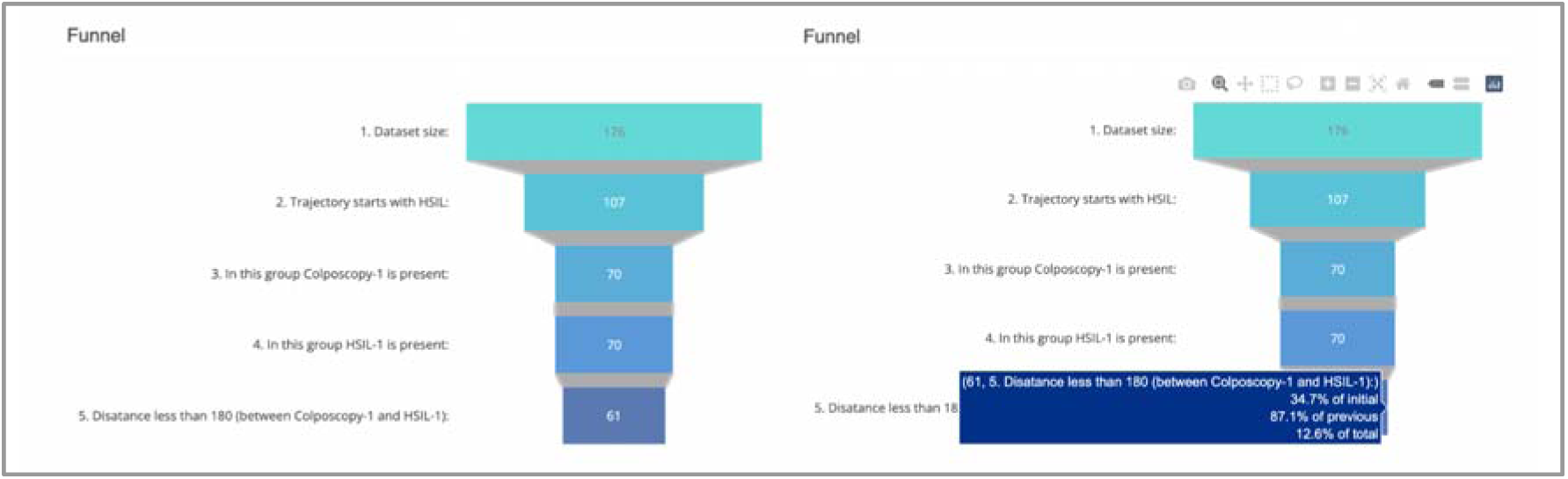
Funnel view describing the data drill-down from the initial dataset to the filtered-out results and interactively showing the percentages between filtered levels in the funnel.

### 3.5 Tabulated views

The plots provide a visual insight into the trajectories from the data. In addition to the visualizations, the package generates several detailed data frames that can be browsed and used to filter specific data. This can provide deeper insight for additional questions like how many pathways are presented in some particular sequence, no matter the level it appears on. Or to study the frequencies of unique pathways within the input data. For example, in our case study, 61 unique sequences were presented, and the most frequent one was “Colposcopy-PAP,” with 15.34% of cases (Figure 6).

**Figure 6.**
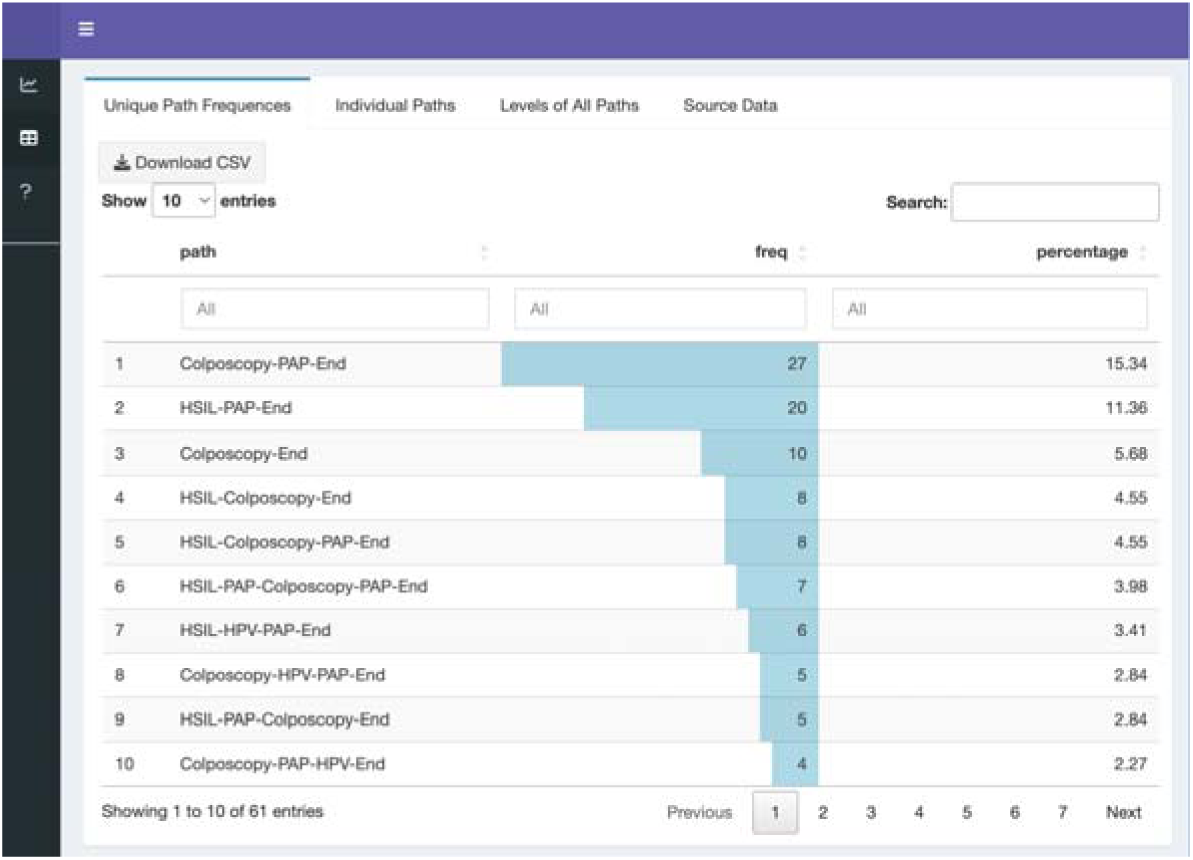
Tables generated from the input data.

## 4 Discussion

We provided a thorough overview and a practical use case of a new user-friendly and interactive R package for visualizing treatment trajectories. This package complements existing solutions by providing a clear overview of the trajectories following or preceding the event under observation and the time gaps between the events. The interactivity of the dashboard allows the re-select and re-align of parts of the initial cohorts to provide a better overview of the data. At the same time, the tabulated view supports the identification of the most frequent trajectories and quantifying specific sequences within unique trajectories.

In recent years, there has been a rapid increase in the number of health databases that have transferred their data to OMOP CDM [13, 14]. In parallel, more and more open-source software to analyze and visualize such data has become available. However, the development of visualization tools for treatment trajectories is lagging. For example, in the ATLAS tool [14], cohort pathways can be visualized with sunburst plots. More recently, R package TreatmentPatterns [15] was introduced where, in addition to sunburst plots, Sankey diagrams are provided to visualize the treatment pathways together with heatmaps to visualize the treatment duration. Our package complements the existing solutions by adding the interactivity to zoom into the specific selection of observed treatment trajectories and adding the time dimension to visualize the periods of treatment or the gaps in between and comparing the periods before or after a specific treatment by aligning the visual trajectories. Another strength of our package is that it relies on standard OMOP infrastructure to define events and communicate with the database; thus, it can be used on any data source in OMOP CDM format. In addition, it works with a more simplistic CSV file prepared for the package input, which expands the use of our package.

Visualizing the real-life treatment trajectories is a great challenge due to the complexity of the data. To draw any conclusions, the patterns have to be simplified to some extent, while on the other hand, oversimplification will lose essential details. Thus, the visualization of treatment trajectories must strike a balance between the two extremes. There is some simplification present also in our package as we need to have one specific event or state defined for a time period, which means that we have to eliminate overlapping events and choose one as a priority when preparing the data with the Cohort2Trajectory package. This might result in omitting a test and only having the test result present in the data (and on the visualization) or vice versa. As a result, there might be a portion of trajectories skewed. The priorities must be defined during the data processing. When preparing the data with the Cohort2Trajectory package, the overlapping states can also be merged to create a new state, but this will make the output of the visualization more busy and difficult to read. Therefore, merging states is more recommended when the number of states is small.

Interactive visualizations, which allow the user to explore various data elements and facets simultaneously, help to simplify the interpretation of the data [2, 3, 4]. Therefore, we have added the possibility of selecting interesting pathways from the sunburst plot and exploring them in more detailed plots.

Using the data tables generated from the source data set can also be beneficial. On the Tables tab, the “Unique Path Frequences” table lists all unique sequences with their frequency and corresponding percentage within the source data. The “Individual Paths” table lists each sequence, enabling the filtering out of all specific event orders independent of the levels at which they occur.

### 4.1 Limitations

The package will process the data set for visualization purposes, and depending on the size and level of pathway complicity, the generation of the first plots might take some time, which the loaders will indicate. Visually limiting will be using datasets where the user would like to look at thousands of patients in detail. The limitation comes from drawing out individual paths that will take up the screen space. This will be addressed in future work by combining information on similar subjects. Still, for now, the package is functional for tens of thousands of subjects when the user drills down via sunburst to the specific groups by their trajectory similarities.

## 5 Conclusion

Our package is a valuable tool for several stakeholders. It is a convenient tool for identifying adherence to the treatment guidelines and detecting all other trajectories and the prevalence of different treatment combinations. In essence, it is a tool to support the understanding of the data and perform exploratory analysis. Thus, the information provided by the package can be used for several purposes, such as generating hypotheses, communicating, designing targeted interventions, and evaluating healthcare services. Therefore, the information from treatment trajectories can be used to develop health policies, make the healthcare system more effective, and provide better care for patients.

In conclusion, we demonstrated how, by going beyond event sequences and exploring the temporal patterns of the sequences, the researcher can better understand the underlying data, spot problems with analysis setup, and generate novel hypotheses. The example study illustrated discovering the adherence to treatment guidelines based on a specific test result.

## Author contributions

All authors participated in the revision process and have approved the submitted version. *Conceptualization* – Maarja Pajusalu, Raivo Kolde. *Data curation* – Maarja Pajusalu, Markus Haug, Marek Oja, Sirli Tamm, Kerli Mooses, Raivo Kolde. Methodology – Maarja Pajusalu, Raivo Kolde. Project administration – Maarja Pajusalu, Raivo Kolde. Resources – Maarja Pajusalu, Raivo Kolde. Software – Maarja Pajusalu, Raivo Kolde. Supervision – Raivo Kolde. Validation – Maarja Pajusalu, Raivo Kolde, Visualization – Maarja Pajusalu, Raivo Kolde. Writing – original draft – Maarja Pajusalu. Writing – Maarja Pajusalu, Raivo Kolde, Kerli Mooses.

## Ethics statement

The study was approved by the Research Ethics Committee of the University of Tartu (300/T-23) and the Estonian Committee on Bioethics and Human Research (1.1-12/653) and the requirement for informed consent was waived.

## Data Availability

There are legal restrictions on sharing de-identified data. According to legislative regulation and data protection law in Estonia, the authors cannot publicly release the data received from the health data registers in Estonia.

https://github.com/HealthInformaticsUT/TrajectoryViz

## Abbreviations

CDM: common data model
OHDSI: Observational Health Data Sciences and Informatics
OMOP: Observational Medical Outcomes Partnership
RWD: real-world data
EHR: electronic health records
HSIL: high-grade squamous intraepithelial lesions

